# Assessing the Effect of Selective Serotonin Reuptake Inhibitors in the Prevention of Post-Acute Sequelae of COVID-19

**DOI:** 10.1101/2022.11.09.22282142

**Authors:** Hythem Sidky, David K. Sahner, Andrew T. Girvin, Nathan Hotaling, Sam G. Michael, Ken Gersing, the N3C consortium

## Abstract

**Importance:** Post-acute sequelae of COVID-19 (PASC) produce significant morbidity, prompting evaluation of interventions that might lower risk. Selective serotonin reuptake inhibitors (SSRIs) potentially could modulate risk of PASC via their central, hypothesized immunomodulatory, and/or antiplatelet properties and therefore may be postulated to be of benefit in patients with PASC, although clinical trial data are lacking.

**Objectives:** The main objective was to evaluate whether SSRIs with agonist activity at the sigma-1 receptor lower the risk of PASC, since agonism at this receptor may serve as a mechanism by which SSRIs attenuate an inflammatory response. A secondary objective was to determine whether potential benefit could be traced to sigma-1 agonism by evaluating the risk of PASC among recipients of SSRIs that are not S1R agonists.

**Design:** Retrospective study leveraging real-world clinical data within the National COVID Cohort Collaborative (N3C), a large centralized multi-institutional de-identified EHR database. Presumed PASC was defined based on a computable PASC phenotype trained on the U09.9 ICD-10 diagnosis code to more comprehensively identify patients likely to have the condition, since the ICD code has come into wide-spread use only recently.

**Setting:** Population-based study at US medical centers.

**Participants:** Adults (≥ 18 years of age) with a confirmed COVID-19 diagnosis date between October 1, 2021 and April 7, 2022 and at least one follow up visit 45 days post-diagnosis. Of the 17 933 patients identified, 2021 were exposed at baseline to a S1R agonist SSRI, 1328 to a non-S1R agonist SSRI, and 14 584 to neither.

**Exposures:** Exposure at baseline (at or prior to COVID-19 diagnosis) to an SSRI with documented or presumed agonist activity at the S1R (fluvoxamine, fluoxetine, escitalopram, or citalopram), an SSRI without agonist activity at S1R (sertraline, an antagonist, or paroxetine, which does not appreciably bind to the S1R), or none of these agents.

**Main Outcome and Measurement:** Development of PASC based on a previously validated XGBoost-trained algorithm. Using inverse probability weighting and Poisson regression, relative risk (RR) of PASC was assessed.

**Results:** A 26% reduction in the RR of PASC (0.74 [95% CI, 0.63-0.88]; P = 5 × 10^−4^) was seen among patients who received an S1R agonist SSRI compared to SSRI unexposed patients and a 25% reduction in the RR of PASC was seen among those receiving an SSRI without S1R agonist activity (0.75 [95% CI, 0.62 - 0.90]; P = 0.003) compared to SSRI unexposed patients.

**Conclusions and Relevance:** SSRIs with and without reported agonist activity at the S1R were associated with a significant decrease in the risk of PASC. Future prospective studies are warranted.

**Key points:** *Question:* Do Selective Serotonin Reuptake Inhibitors with and without agonist activity at the sigma-1 receptor (S1R) prevent Post-Acute Sequelae of COVID-19?

*Findings:* In this retrospective study leveraging real-world clinical data that included 17 933 patients, a 28% reduction in risk of PASC was observed for S1R agonist SSRIs and a 25% reduction in risk of PASC was observed for non-S1R agonist SSRIs, both versus controls, using a computable phenotype to define PASC.

*Meaning:* SSRIs may play a role in managing the long term disease burden of COVID-19. Future prospective studies are warranted to confirm these findings and evaluate potential mechanisms of action.

## Introduction

### Post-Acute Sequelae of SARS-CoV-2

Post-Acute Sequelae of SARS-CoV-2 Infection (PASC) or long COVID has been estimated by the World Health Organization to afflict between ∼10-20% of patients with COVID-19, although estimates vary considerably based on methodology.^1^ At least one relatively large EHR-based study suggested that approximately one-third of patients may have symptoms compatible with PASC 3-6 months after diagnosis of COVID-19, with a higher risk among females and those with more severe disease.^2^ Symptoms of PASC are generally nonspecific and include dyspnea, cough, chest pain, headache, arthralgia, myalgia, fatigue, post-exertional malaise or poor endurance, fever, “brain fog” or cognitive impairment, paresthesia, insomnia, anosmia, dysgeusia, mood alterations, palpitations or tachycardia (which may be postural/orthostatic), lightheadedness, abdominal pain, diarrhea, menstrual irregularities and rash.^3–5^ These symptoms may substantially impair function, persist for an extended duration, and fluctuate over time.^2,6^ At present, there is no specific recommended treatment for PASC other than supportive care and rehabilitation,^7^ although some preliminary observational and cross-sectional data suggest that SARS-CoV-2 vaccination, either before or shortly after infection, may attenuate the risk of PASC.^8–10^

### Pathogenesis of PASC

The characterization of PASC, its potentially variable phenotypes and possible pathophysiological underpinnings, remains at a relatively early stage, although insights have been provided by a number of studies. Hypotheses that have been adduced have focused primarily on immune dysregulation and/or auto-immunity (discussed below), but other speculative explanations have also been put forth, including redox imbalance and/or mitochondrial dysfunction,^11–13^ persistence of virus in sanctuary sites,^14,15^ and formation of fibrinolysis-resistant amyloid fibrin microclots.^16^

Most work attempting to clarify the pathogenesis of PASC has centered on a potentially aberrant immune response primarily because little to no evidence exists of productive viral replication in the vast majority of patients. Although viral RNA has been detected in various tissues at autopsy after extended follow-up^17^ and shedding of virus in the stool may persist after respiratory shedding has ceased, prolonged shedding of virus is unusual, with notable exceptions including reports in immunocompromised patients.^18,19^ Swank and colleagues^15^ customized an existing assay in order to detect spike and/or nucleocapsid antigen in the blood of 31 patients with PASC and found evidence for prolonged antigenemia, but, importantly, discordance was noted between long-term expression of spike (60% of patients) and nucleocapsid (a single patient). This raises the possibility of subgenomic integration of reverse transcribed viral genetic material into host DNA leading to prolonged antigen production without productive viral infection, which is supported by prior *in vitro* work,^20^ since one would also expect to see nucleocapsid antigen in the context of productive viral infection. To our knowledge, there have been no reports of virus cultured from the blood of any patient with PASC.

Su and colleagues^21^ extensively characterized a primary cohort of over 200 SARS-CoV-2-infected inpatients and outpatients followed longitudinally for up to 2-3 months versus matched controls. In addition, a separate cohort of 100 COVID-19 inpatients and outpatients was asked to return at 60 or 90 days of follow-up and 33 COVID-19 patients from a third cohort also provided plasma samples to validate some of the findings. Significant associations were variably seen between certain late PASC manifestations and autoantibodies during acute infection directed at IFNα-2, U1-snRNP, and LL/SS-B, but given the number of comparisons and limited sample size, it is difficult to draw definitive conclusions from this observation.

Of note, 44% of patients in the study by Su and colleagues had autoantibodies at the extended follow-up visit but 56% of patients had class-switched autoantibodies during the acute infection, suggesting that antibodies antedated COVID-19 infection. The vast majority of patients with autoantibodies had no prior diagnosis of an auto-immune condition. Patients with elevated plasma IFN-γ during acute infection had elevated autoantibodies directed at IFNα-2 at extended follow-up, and those with increased plasma levels of the chemokine CXCL9 exhibited elevated autoantibody to P1 at delayed follow-up (P < 0.001). There was an anticorrelation between virus-specific antibody levels and autoantibodies (antinuclear and anti-IFNα-2). CMV-specific CD8+ T cells were seen at late follow-up in patients with gastrointestinal PASC symptoms, leading to the hypothesis that bystander activation of these cells may be seen in GI PASC. Several immune phenotypes at delayed follow-up were variably associated with different PASC symptoms, including myeloid-derived suppressor cells, memory-like NK cells, and innate immune activation, whereas activated Treg cells during acute infection seemed to augur PASC.

Vijayakumar et al. identified an immunoproteomic signature in patients with respiratory disease following acute COVID-19, particularly using BAL (airway) samples. An increase in activated CD8+ T cells in bronchoalveolar lavage fluid was associated with a reduction in forced vital capacity (FVC).^22^ Chemokines that recruit T cells and NK cells were also highly associated with epithelial damage markers. In addition, increased memory B cells in airways was linked to reduction in FVC or TCLO, or imaging abnormalities.

Markers of immune activation and inflammation among patients with PASC were evaluated by Peluso et al.^23^ Trends in these data revealed that patients with PASC were more likely to be female (61.6%) and to have a history of autoimmune disease (11%) versus those who had recovered from COVID-19 without PASC (2.1%). Significantly higher levels of TNF-α and IP-10, and a trend toward higher levels of IL-6, were seen during early recovery among those patients who developed PASC. At the late recovery time point, IL-6 levels were significantly higher among patients with PASC. In another study, compared with healthy controls patients with PASC had higher plasma levels of CCL5/RANTES, IL-2, IL-4, CCL3, IL-6, IL-10, IFN-γ, and VEGF; decreased T regulatory cells and B cell elevations were also seen.^24^

In aggregate, the data above suggest that immune dysregulation, associated with increased systemic and/or pulmonary levels of chemokines, pro-inflammatory cytokines, and specific lymphocyte phenotypes, may be responsible for the pathogenesis of PASC, although further study is needed, particularly of larger cohorts of patients followed prospectively, with intensive evaluation of both plasma and cell-based biomarkers. Immune dysregulation may be associated with autoantibodies, and the potential role of cross-reactive antibodies directed at host targets cannot be excluded, but early published data suggest that these autoantibodies may antedate COVID-19, possibly identifying a subset of patients predisposed to develop PASC.

### SSRIs as Immunomodulatory Agents in COVID-19

In contrast to direct antiviral effects, existing data suggest that SSRIs may harbor potential as *immunomodulators* that favorably affect the clinical course of COVID-19 and/or the risk of PASC. Abundant in vitro, animal model, and, to some extent, clinical data suggest SSRIs have immunomodulatory properties, primarily immunosuppressive activity.^25–30^ In interpreting these data, attention must be paid, however, to dose, concentration, activation status of lymphocytes, experimental design and the underlying disease/model. Doses and concentrations in preclinical studies are often higher than those in humans. For example, concentrations at which lymphocyte proliferation is inhibited by SSRIs are typically ≥ 1 μM, higher than those typically achieved in the clinic. Sertraline, however, at concentrations of 0.01 and 1 µM decreases the IFN/IL-10 ratio in the supernatant of mitogen-stimulated whole blood. SSRIs deplete platelet serotonin and, in mice, platelet-derived serotonin appears to promote neutrophil endothelial adhesion and inflammation-triggered extravasation of neutrophils into tissue. Several other mechanisms have been proposed to explain the immunomodulatory effects of SSRIs. At first, inhibition of the serotonin transporter protein, resulting in increased extracellular concentrations of 5-HT, was thought to be responsible for the observed effects of SSRIs on lymphocytes, but multiple lines of experimental evidence contravene this hypothesis.^26^ The impact of SSRIs on several signal transduction pathways has been posited as the basis for diminished lymphocyte proliferation and apoptosis; these effects include activation of PKA through induction of cAMP, inhibition of the translocation of PKC to the cell surface, and reduction in calcium influx.^26^ However, reduced lymphocyte proliferation and apoptosis are generally not expected at typical clinical concentrations of commonly used SSRIs.

Recently, attention has focused on the observation that several SSRIs bind with moderate to high affinity to the sigma-1 receptor (SIR), and it has been suggested that ligands for this receptor may modulate the immune system. The S1R is a ubiquitous endoplasmic reticulum (ER)-resident chaperone protein that associates with IRE1 in the context of ER stress, leading to regulation of the production of inflammatory cytokines. IRE1 is necessary for cytokine production, presumably through XBP1-mediated transactivation of IL-6 and TNF-α. Fluvoxamine, which exhibits the highest affinity for the S1R among antidepressants and exerts agonistic activity, has been shown to produce anti-inflammatory effects in a variety of cell models.^31^ Rosen et al.^32^ showed that S1R KO mice experienced increased mortality in two sub-lethal models of sepsis, and that WT mice treated with fluvoxamine were protected from mortality and had lower IL-6 levels in a lethal model of sepsis. Data suggest that, at least for some SSRIs, typical clinical concentrations may result in appreciable binding at the S1R (see Table S1 in Supplementary Appendix). In particular, based on published data, we have calculated approximate ratios of free (unbound) typical clinical concentrations to the dissociation constant at the sigma-1 receptor of 2-8.5 for fluvoxamine, 0.2 - 0.6 for citalopram, 0.2 to 0.4 for escitalopram, 0.1 to 0.5 for fluoxetine and 0.1 or less for sertraline. Of note, fluvoxamine, fluoxetine, escitalopram, and, presumably citalopram are agonists at the S1R whereas sertraline is an antagonist; paroxetine is not expected to bind appreciably to the S1R.^33^

Whatever the mechanistic basis might be, some clinical data suggest that cytokine levels in the clinic may be affected by SSRIs. In a large meta-analysis of largely uncontrolled data from longitudinal studies of depressed patients treated pharmacologically, IL-6 levels fell significantly irrespective of treatment response.^34^ This analysis was not stratified by treatment, but, in another meta-analysis of pharmacologic treatment for depression that partially overlapped with the study above, changes in cytokine levels were also assessed by drug class.^35^ Overall, treatment reduced levels of IL-1β. Selective serotonin reuptake inhibitors were also found to potentially affect levels of IL-6 and TNF-α, but other antidepressants in this meta-analysis did not appear to reduce measured cytokine levels. In a small randomized controlled study decreases in IL-1β were seen among depressed patients treated with fluoxetine or acupuncture vs. placebo,^36^ but only 68/95 subjects contributed baseline samples, and cytokine data were available at follow-up for only 72 subjects. There was no apparent impact of fluoxetine on TNF-α or Th2 cytokine levels. Other data suggest that other forms of treatment for depression may influence cytokine levels (Dahl et al., 2014; Brunoni et al., 2014).^37,38^ Thus, the vast majority of the available clinical data suggesting SSRIs may reduce cytokine levels derive from uncontrolled studies, and the paucity of prospective randomized controlled data in which cytokine data have been consistently measured make it difficult to determine if this effect is specific to SSRIs in the context of treatment for depression.

There have been several published randomized controlled double-blind trials of fluvoxamine in outpatients with acute COVID-19, two of which have demonstrated preliminary but non-conclusive evidence of benefit ^39,40^ in the absence of any evidence of antiviral effect.^40^ Two other prospective studies did not demonstrate an impact, although this may have been related, in part, to the choice of dose of fluvoxamine.^41,42^ Although all of these studies have limitations, we hypothesize that these results are consistent with an immunological basis for protection against the cytokine storm elicited in some patients by SARS-CoV-2,^43^ suggesting further study is needed. In view of all of the above findings, suggesting SSRIs (a) exert immunomodulatory effects, possibly through sigma-1 receptor (S1R) binding, and (b) might enhance clinical outcomes in patients with acute COVID-19, we hypothesized that chronically administered SSRIs binding to the S1R with relatively high affinity, and exhibiting sigma-1 receptor agonism, might dampen a pro-inflammatory immune response and thereby reduce the risk of PASC. As noted above, some data suggest that ongoing immune dysregulation may be responsible for the pathogenesis of PASC. We therefore evaluated, in this retrospective study relying upon a dataset including de-identified EHR and PPRL-linked ancillary data, the risk of PASC among recipients of commonly prescribed SSRIs. In particular, we studied the effects of SSRIs with (a) agonist activity at the S1R, (b) antagonist activity at the S1R, or (c) no meaningful binding at the S1R in an effort to discern whether SIR agonism played a role in any observed beneficial effect.

## Methods

### Setting and population

We used data from the National COVID Cohort Collaborative (N3C), a centralized real-world repository of de-identified electronic health records supported by the National Institutes of Health (NIH). N3C includes detailed information on clinical encounters including procedures, diagnoses, ordered and administered medications, demographic data, vitals, and lab orders and results. Records in N3C are aggregated across participating clinical organizations in the United States, known as data partners, harmonized using the Observational Medical Outcomes Partnership (OMOP) data model, and subjected to quality review and checks. This research was possible because of the patients whose information is included within the data and the organizations (https://ncats.nih.gov/n3c/resources/data-contribution/data-transfer-agreement-signatories) and scientists who have contributed to the on-going development of this community resource. The N3C data transfer to NCATS is performed under a Johns Hopkins University Reliance Protocol #IRB00249128 or individual site agreements with the NIH. The N3C Data Enclave is managed under the authority of the NIH; information can be found at https://ncats.nih.goc/n3c/resources. The content is solely the responsibility of the authors and does not necessarily represent the official views of the National Institutes of Health or the N3C program. Use of N3C data for this study does not involve human subjects (45 CFR 46.102) as determined by the NIH Office of IRB Operations.

We included adult patients (≥ 18 years) with either RT-PCR or antigen (AG) confirmed SARS-CoV-2 infection or a recorded U07.1 diagnosis for COVID-19. Due to the recent availability of the U09.9 diagnosis code for PASC on which our computable phenotype was trained (see below), only patients with an index date, defined as the earliest record of infection or diagnosis, on or after October 1, 2021 through April 7, 2022 were included. This would mitigate the potential lack of generalizability of the algorithm over time and also limit the analysis to the inclusion of subjects with either Delta or Omicron infection. Patients were required to have at least one interaction with the participating health care system prior to and following the index diagnosis of COVID-19. The analysis was restricted to participating N3C sites with vaccination rates matching those reported by the CDC for the corresponding geographic region, as vaccination may affect the risk of PASC and was considered a covariate. To allow enough time for collection of data relevant to a PASC diagnosis, we included only patients with at least 90 days between their index diagnosis date and the data extraction date. We excluded patients with missing gender data and those who died during COVID-19-related hospitalization or within 45 days of their index date.

### Exposures

Three groups were defined: (1) patients with documented baseline exposure to least one SSRI with established or presumed agonist activity at the S1R (fluvoxamine, fluoxetine, escitalopram, or citalopram), (2) patients with documented baseline exposure to at least one SSRI without presumed agonist activity at S1R (sertraline, an antagonist, or paroxetine, which does not appreciably bind to the S1R), and (3) control patients with no documented baseline exposure to either (1) or (2). Baseline use of a drug was defined as recorded at or prior to the COVID-19 index diagnosis date. Records in N3C date back to January 1, 2018 for patients who had been followed for the longest period of time. Patients with documented baseline exposure to both (1) and (2) were excluded. Treatment comparisons were performed between patients in the exposure group (1) versus controls and the exposure group (2) versus controls. To further interrogate the effect of S1R agonism as a mechanism of action, we conducted a secondary analysis with non-S1R agonist SSRI exposure as a comparator to S1R agonist SSRI exposure.

### Outcomes

The primary outcome consisted of a presumptive diagnosis of PASC using an XGBoost-based machine learning algorithm trained on patients assigned the U09.9 (PASC) diagnosis code.^44^ This surrogate was chosen because the U09.9 code is of relatively recent vintage (established in October of 2021) and had not yet been rapidly and fully embraced by all participating healthcare organizations. Neither vaccination nor the SSRIs under study were included as features in this predictive model. The prediction threshold for the model was chosen to be 0.53 (see Figure 1), which corresponds to 15% of the total patient population receiving a PASC diagnosis. Rates of PASC reported in the literature have varied widely, but the likely rate is generally believed to reside within a range of approximately 10-20%.^1^ In view of this, a supplementary analysis was performed using a lower and higher score threshold yielding a rate of 10% and 20% respectively.

**Figure 1.**
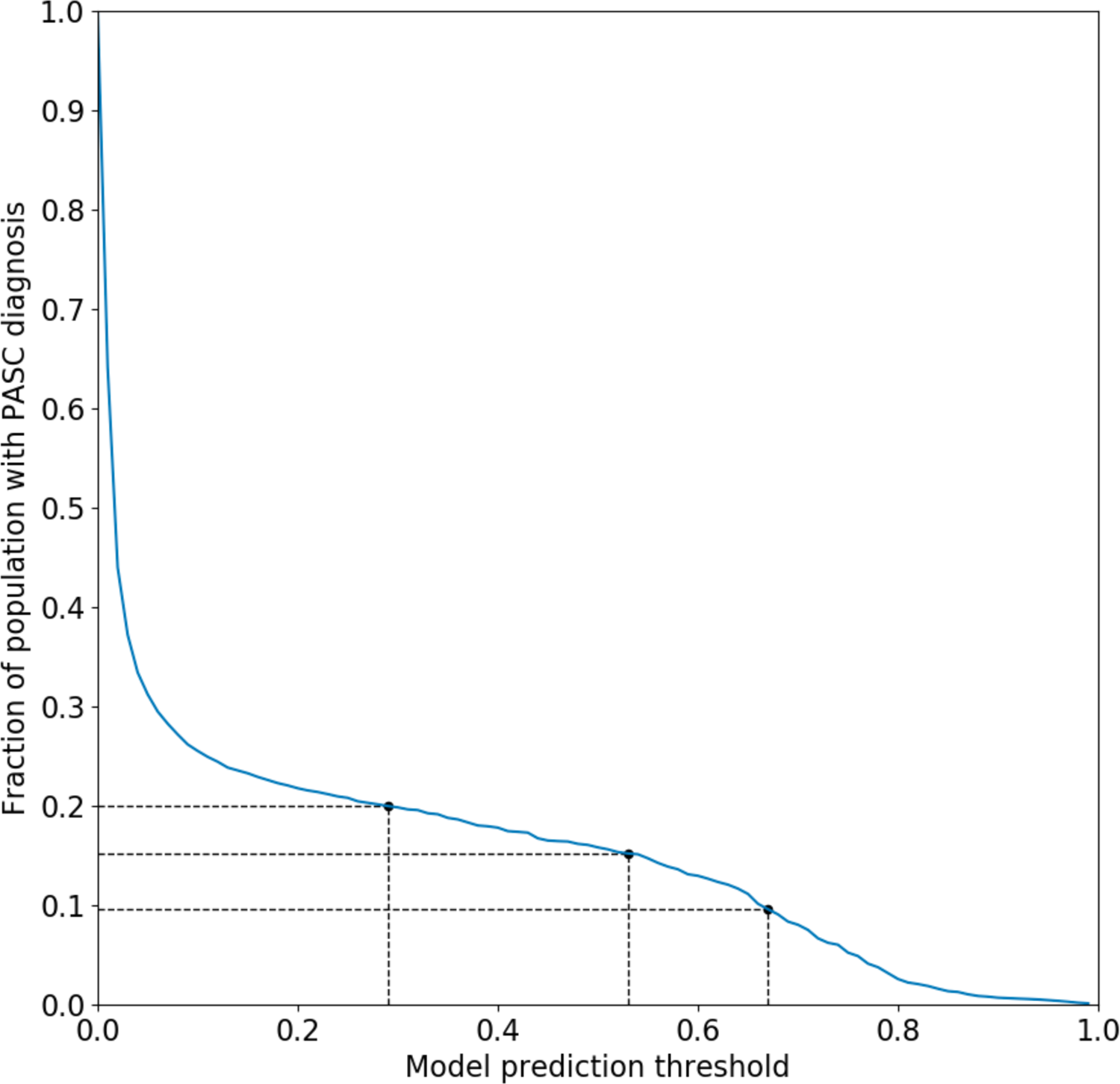
Fraction of patient population receiving a PASC diagnosis as a function of predictive model probability threshold. A cutoff of 0.53 was chosen for the main analysis which corresponds to 15% of the patient population. Cutoffs of 0.29 and 0.67 are also shown, corresponding to 10% and 20% of the population respectively.

### Statistical analysis

For the primary analyses we first compared the risk of PASC among patients receiving a moderate to high-affinity S1R agonist SSRI (fluvoxamine, fluoxetine, escitalopram, or citalopram) at baseline to patients not receiving any SSRI. Second, we compared the risk of PASC among patients receiving an SSRI with no high-affinity S1R agonism (sertraline, paroxetine) at baseline to patients not receiving any SSRI. Note that citalopram was assumed to be an agonist because escitalopram (the S-enantiomer of citalopram) is a known S1R agonist. Relative risks (RRs) and 95% CIs were estimated using Poisson regression with robust standard errors. Inverse probability weighting (IPW) as implemented in the R WeightIt package, version 0.9.0, was used to adjust for confounding. The individual exposure propensity scores were calculated using multivariate logistic regression and included age, sex, body mass index (BMI), race and ethnicity, baseline comorbidities (myocardial infarction, congestive heart failure, cerebrovascular disease, dementia, chronic lung disease, uncomplicated type II diabetes, complicated type II diabetes, kidney disease, liver disease, HIV infection, and cancer), record of post-infection hospitalization, use of approved immunomodulatory therapy for acute COVID-19 (dexamethasone, baricitinib, tocilizumab), baseline immunotherapy treatment (etanrecept, infliximab, adalimumab, certolizumab pegol, golimumab, sarilumab, tocilizumab, baricitinib, tofacitinib, or upadacitinib), baseline exposure to other ligands exhibiting S1R binding with moderate to high affinity defined as a dissociation constant of less than 250 nanomolar (trifluoperazine, pimozide, fluphenazine, chlorpromazine, perphenazine, haloperidol, pentazocine, progesterone, methamphetamine, hydroxychloroquine, dextromethorphan, clemastine, hydroxyzine, tamoxifen), any baseline SNRI medication (desvenlafaxine, duloxetine, levomilnacipran, venlafaxine), baseline bupropion exposure, any baseline tricyclic antidepressant medication (clomipramine, amoxapine, amitriptyline, desipramine, nortriptyline), any baseline benzodiazepine medication (alprazolam, chlordiazepoxide, diazepam, lorazepam), any baseline antipsychotic medication (risperidone, quetiapine, ziprasidone, aripiprazole, aripiprazole lauroxil, clozapine), and any record of COVID-19 vaccination. For BMI, which was the only variable with missing data, a missing data indicator was created. The estimated probabilities were used to calculate stabilized inverse probability weights and used to weight each patient’s contribution to the Poisson regression model. Covariate balance before and after inverse probability weighting was assessed by calculating absolute standardized mean differences (SMDs) and comparing the distributions of propensity scores for exposed and unexposed groups, which were generated using the R Cobalt package, version 4.2.2.

We performed additional analyses using the methods detailed above for the primary analysis. To isolate and determine the significance of S1R agonism in reducing the risk of long-COVID, we used non-S1R agonist SSRIs (sertraline and paroxetine) as a comparator to high-affinity S1R agonist SSRIs. Sertraline is an antagonist at the S1R and paroxetine, an SSRI for which agonist versus antagonist activity is not established, is not expected to meaningfully bind to the S1R given its high dissociation constant.^33^ The effect of both SSRI classes across sex and COVID-19 vaccination status (record of any COVID-19 vaccination vs. unvaccinated) subgroups was also assessed. As a sensitivity analysis, we repeated our primary analysis with lower and higher probability thresholds for the PASC computable phenotype ML model of 0.67 and 0.29, which correspond to 10% and 20% of the population respectively, to establish the robustness of the results to the long-COVID diagnosis assignments. Finally, to assess the impact of increased specificity to recent ongoing SSRI treatment prior to infection, we limited the exposure definition to include only patients with a record of exposure within 180 days of the index COVID-19 diagnosis date.

Data extraction was carried out in the N3C enclave using Spark SQL 3.2.1 and Python 3.6.7, and all statistical analysis was done using R version 3.5.

## Results

There were 17 933 eligible patients identified, with a total of 2021 patients exposed to S1R agonist SSRIs at baseline, 1328 exposed to non-S1R agonist SSRIs, and 14 584 unexposed patients (Figure 2). Both the S1R and non-S1R agonist SSRI exposed patients were more female compared with unexposed patients (1546 [76%] and 975 [73%] vs 8943 [61%]), slightly younger (median [IQR] age: 49 [36, 62] and 47 [34, 62] vs 50 [36, 64]), and more White non-Hispanic (1579 [78%] and 987 [74%] vs 9860 [68%]). Exposed patients generally had more comorbidities (congestive heart failure, 209 [10%] and 153 [12%] vs 1181 [8.1%]; cerebrovascular disease, 148 [7.3%] and 92 [6.9%] vs 641 [4.4%]; chronic lung disease, 583 [29%] and 370 [28%] vs 2967 [20%]; uncomplicated type II diabetes, 451 [22%] and 315 [24%] vs 2793 [19%]; cancer, 241 [12%] and 178 [13%] vs 1621 [11%]); exposed patients were also more likely to have been exposed to other high S1R affinity ligands at baseline (548 [27%] and 362 [27%] vs 1931 [13%]), and more likely to have received other classes of psychiatric medication at baseline (bupropion, 372 [18%] and 172 [13%] vs 586 [4.0%]; benzodiazepines, 706 [35%] and 395 [30%] vs 2113 [14%]; tricyclic antidepressants, 132 [6.5%] and 83 [6.2%] vs 392 [2.7%]; antipsychotics, 178 [8.8%] and 103 [7.8%] vs 298 [2.0%]; SNRIs, 257 [13%] and 165 [12%] vs 987 [6.8%]). Exposed patients were more likely to have received a COVID-19 vaccine (1353 [67%] and 904 [68%] vs 8744 [60%]). COVID-19 related hospitalization was similar across all three groups (287 [14%] and 193 [15%] vs 2308 [16%]) as was baseline immunotherapy (34 [1.7%] and 24 [1.8%] vs 244 [1.7%]) and COVID-19 immunotherapy (37 [1.8%] and 30 [2.3%] vs 372 [2.6%]) but unexposed patients, unadjusted, were more likely to develop long-COVID (233 [12%] and 155 [12%] vs 2352 [16%]). Complete cohort characteristics are shown in Table 1.

**Table 1.** Cohort characteristics by SSRI exposure group. Note that small numbers (<20) were obscured to protect patient identities.

| Characteristic | S1R agonist SSRI exposure<br>N = 2021<sup>1</sup> | Non-S1R agonist SSRI exposure<br>N = 1328<sup>1</sup> | Unexposed<br>N = 14 584<sup>1</sup> |
| --- | --- | --- | --- |
| **Sex** |  |  |  |
| Male | 475 (24%) | 353 (27%) | 5641 (39%) |
| Female | 1545 (76%) | 975 (73%) | 8943 (61%) |
| **Age, median (years)** | 49 (36, 62) | 47 (34, 62) | 50 (36, 64) |
| **Age (years)** |  |  |  |
| 18-49 | 1017 (50%) | 701 (53%) | 7006 (48%) |
| ≥70 | 291 (14%) | 214 (16%) | 2332 (16%) |
| 50-59 | 406 (20%) | 237 (18%) | 2765 (19%) |
| 60-69 | 307 (15%) | 176 (13%) | 2481 (17%) |
| **BMI, median (kg/m<sup>2</sup>)** | 33 (28, 39) | 33 (28, 39) | 31 (27, 37) |
| **BMI (kg/m<sup>2</sup>)** |  |  |  |
| <25 | 220 ± 5 (11%) | 150 ± 5 (11%) | 1957 (13%) |
| ≥40 | 500 ± 5 (25%) | 315 ± 5 (24%) | 2518 (17%) |
| 25-29 | 445 ± 5 (22%) | 290 ± 5 (22%) | 3595 (25%) |
| 30-34 | 500 ± 5 (25%) | 335 ± 5 (25%) | 3457 (24%) |
| 35-39.9 | 345 ± 5 (17%) | 230 ± 5 (17%) | 2250 (15%) |
| Missing | < 20 | < 20 | 807 (5.5%) |
| **Race and ethnicity** |  |  |  |
| White Non-Hispanic | 1579 (78%) | 987 (74%) | 9860 (68%) |
| Asian Non-Hispanic | 27 (1.3%) | 20 (1.5%) | 309 (2.1%) |
| Black or African American Non-Hispanic | 279 (14%) | 222 (17%) | 3094 (21%) |
| Hispanic or Latino Any Race | 81 (4.0%) | 70 (5.3%) | 899 (6.2%) |
| Other Non-Hispanic / Unknown | 55 (2.7%) | 29 (2.2%) | 422 (2.9%) |
| **Myocardial infarction** | 88 (4.4%) | 67 (5.0%) | 630 (4.3%) |
| **Congestive heart failure** | 209 (10%) | 153 (12%) | 1181 (8.1%) |
| **Cerebrovascular disease** | 148 (7.3%) | 92 (6.9%) | 641 (4.4%) |
| **Dementia** | 58 (2.9%) | 53 (4.0%) | 245 (1.7%) |
| **Chronic lung disease** | 583 (29%) | 370 (28%) | 2967 (20%) |
| **Type II diabetes, uncomplicated** | 451 (22%) | 315 (24%) | 2793 (19%) |
| **Type II diabetes, complicated** | 266 (13%) | 191 (14%) | 1564 (11%) |
| **Kidney disease** | 255 (13%) | 194 (15%) | 1567 (11%) |
| **Liver disease** | 225 (11%) | 116 (8.7%) | 1009 (6.9%) |
| **HIV infection** | < 20 | < 20 | 82 (0.6%) |
| **Cancer** | 241 (12%) | 178 (13%) | 1621 (11%) |
| **COVID-19 related hospitalization** | 287 (14%) | 193 (15%) | 2308 (16%) |
| **Baseline bupropion use** | 372 (18%) | 172 (13%) | 586 (4.0%) |
| **Baseline benzodiazepine use** | 706 (35%) | 395 (30%) | 2113 (14%) |
| **Baseline tricyclic antidepressant use** | 132 (6.5%) | 83 (6.2%) | 392 (2.7%) |
| **Baseline antipsychotic use** | 178 (8.8%) | 103 (7.8%) | 298 (2.0%) |
| **Baseline SNRI use** | 257 (13%) | 165 (12%) | 987 (6.8%) |
| **Baseline immunotherapy treatment** | 34 (1.7%) | 24 (1.8%) | 244 (1.7%) |
| **Baseline moderate to high affinity S1R ligand use** | 548 (27%) | 362 (27%) | 1931 (13%) |
| **COVID-19 related immunotherapy** | 37 (1.8%) | 30 (2.3%) | 372 (2.6%) |
| **COVID-19 vaccine** | 1353 (67%) | 904 (68%) | 8744 (60%) |
| **Long-COVID** | 233 (12%) | 155 (12%) | 2352 (16%) |

<sup>1</sup> Statistics presented: n (%); median (IQR).

**Figure 2.**
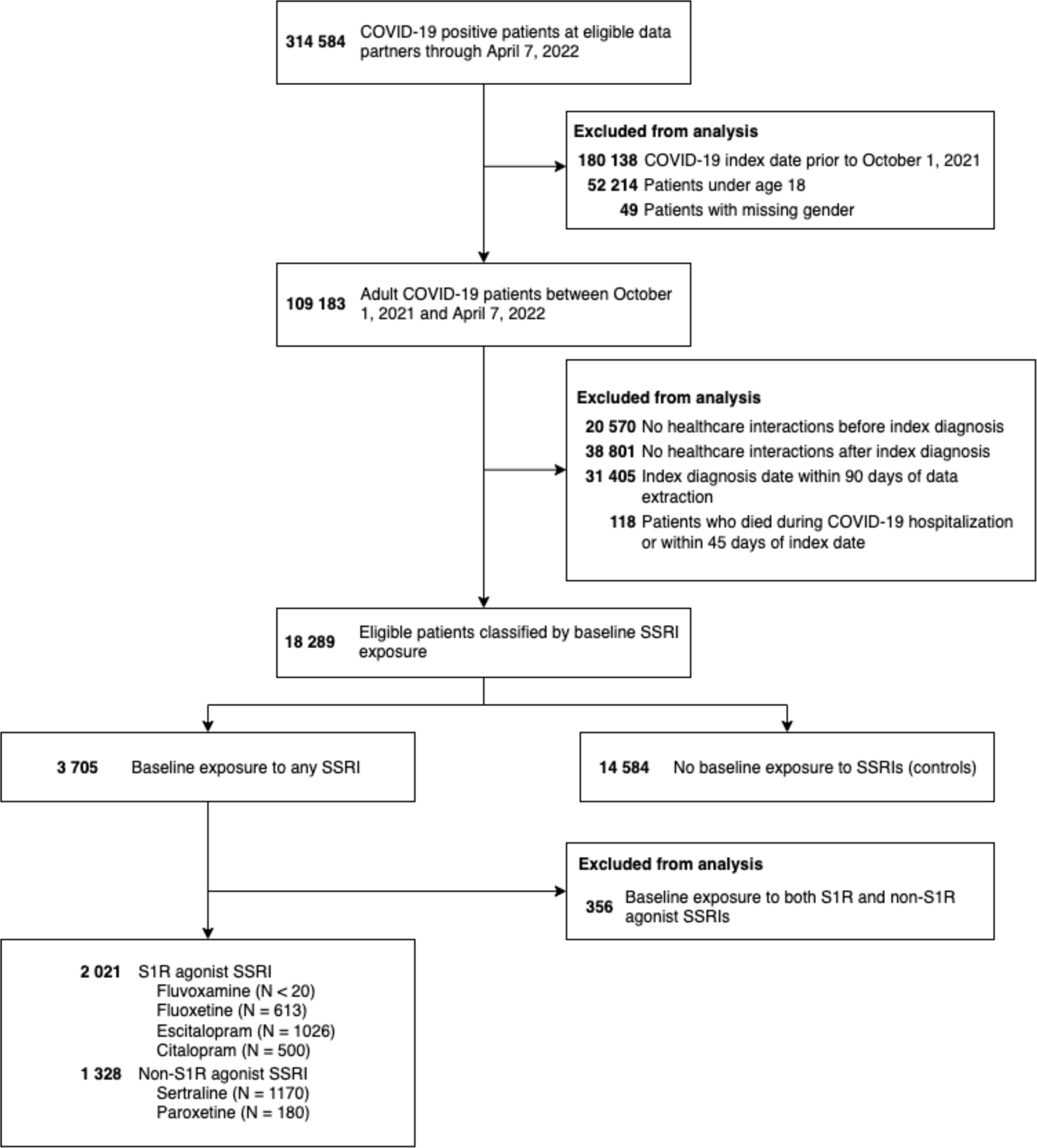
Flow diagram of patient selection. Note that SSRI exposure is non-exclusive and small numbers (<20) were obscured to protect patient identities.

After applying IPW, patient characteristics between exposed and unexposed groups for both S1R agonist and non-S1R agonist SSRIs analyses were adequately balanced, with absolute standardized mean differences (SMDs) under 0.1 for almost all covariates. The only two exceptions were baseline SNRI use and other non-Hispanic or unknown ethnicity for the S1R agonist SSRI analysis - both at 0.1. Figure 3 shows the SMDs for treated and control groups along with the propensity score distributions before and after weighting. Covariate balance for secondary and subgroup analyses were also adequate and are presented in the Supplement (Figures S1 to S11).

**Figure 3.**
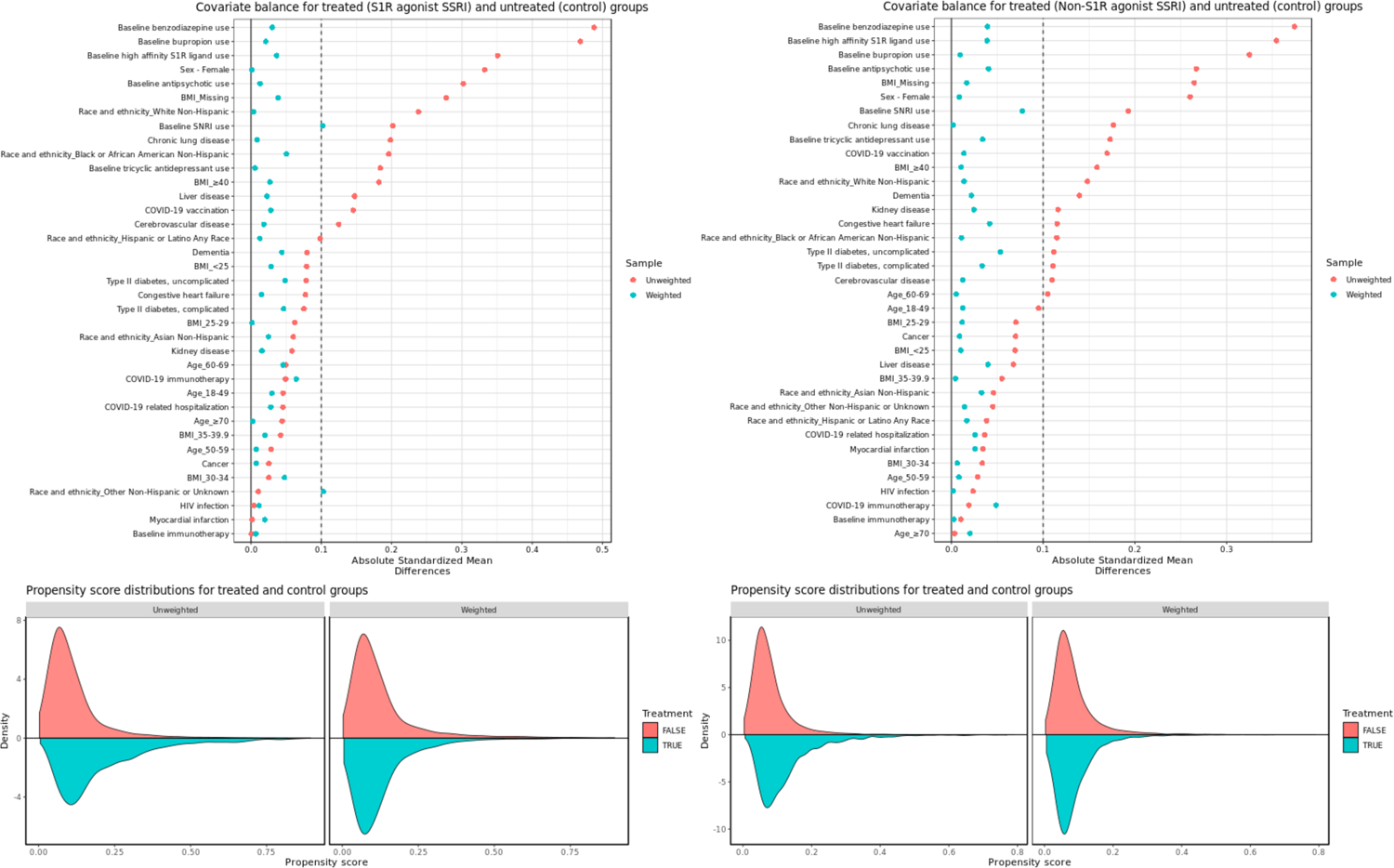
Covariate balance and propensity score distributions before and after weighting for (left) S1R agonist SSRI exposed groups vs control and (right) non-S1R agonist SSRI exposed groups vs control.

In the weighted analysis, there was a 26% reduction in the RR of long-COVID (0.74 [95% CI, 0.63-0.88]; P = 5 × 10^−4^) among patients with baseline S1R agonist SSRI exposure compared to unexposed patients. For patients with baseline non-S1R agonist SSRI exposure compared to unexposed patients, there was a 25% reduction in RR (0.75 [95% CI, 0.62 - 0.90]; P = 0.003). When considering non-S1R agonist SSRIs as a comparator, no statistically significant difference in RR (0.95 [95% CI, 0.78 - 1.1]; P = 0.6) was observed between S1R agonist SSRI recipients and non-S1R agonist SSRIs. Additional analysis showed that the significance of RR reduction among both genders for S1R agonist SSRI exposure was maintained (male: 0.61 [95% CI, 0.43 - 0.85], P = 0.003; female: 0.80 [95% CI, 0.67 - 0.96], P = 0.02) but not for non-S1R agonist SSRIs (male: 0.57 [95% CI, 0.38 - 0.86], P = 0.008; female: 0.88 [95% CI, 0.71 - 1.1], P = 0.3). Subgroup analysis on vaccinated and non-vaccinated patients with S1R agonist SSRI exposure revealed a significant RR reduction for vaccinated patients (0.73 [95% CI, 0.58 - 0.93]; P = 0.01) but not among unvaccinated patients (0.82 [95% CI, 0.64 - 1.1]; P = 0.1) and a lack of significant RR reduction for both vaccinated (0.78 [95% CI, 0.59 - 1.0]; P = 0.09) and unvaccinated patients (0.79 [95% CI, 0.59 - 1.1]; P = 0.1) with non-S1R agonist SSRI exposure.

The results of our primary analyses proved to be fairly robust to the ML model threshold for Long-COVID assignment. At a threshold of 0.29 where 20% of the patient population received a long-COVID diagnosis, we still observed a 17% reduction in the RR of long-COVID (0.83 [95% CI, 0.73 - 0.96]; P = 0.01) among S1R agonist SSRI exposed patients and a 22% reduction in RR of long-COVID (0.78 [95% CI, 0.67 - 0.91]; P = 0.002) among non-S1R agonist SSRI exposed patients, both compared to unexposed patients. At a threshold of 0.67 where 10% of the patient population received a long-COVID diagnosis, RR significance is lost for S1R agonist exposure (0.87 [95% CI, 0.72 - 1.1]; P = 0.2) but maintained for non-S1R agonist exposure versus control (0.77 [95% CI, 0.60 - 0.97]; P = 0.03). Furthermore, the second sensitivity analysis limiting the SSRI exposure window to 180 days prior to the COVID-19 index date yielded a lower RR (0.66 [95% CI, 0.54 - 0.82]; P = 1 × 10^−4^) compared to the primary analysis for the S1R agonist exposure. For the corresponding non-S1R agonist exposure analysis, the RR (0.78 [95% CI, 0.62 - 0.99]; P = 0.04) was similar to the primary analysis. Figure 4 summarizes the RR estimates for all these analyses.

**Figure 4.**
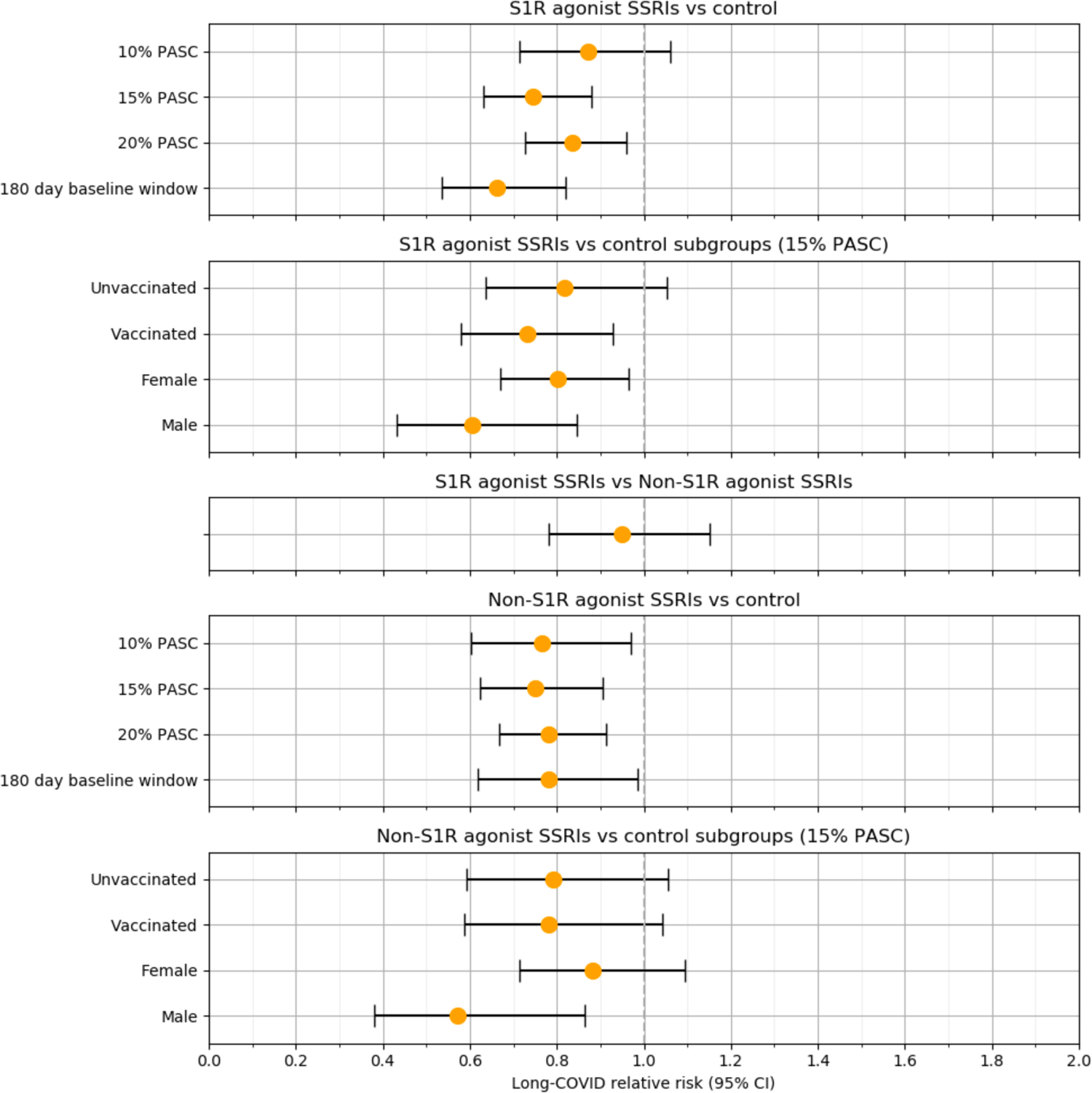
Associations between baseline S1R agonist and non-S1R agonist SSRI exposure and long-COVID for different patient groups and model specifications.

## Discussion

In this multicenter retrospective study, we observed a statistically significant 26% reduction in the RR of long-COVID among patients receiving baseline S1R agonist SSRIs when compared with controls. A similar effect size of 25% reduction in the RR of long-COVID was observed for patients receiving non*-*S1R agonist SSRIs compared to controls. Furthermore, when comparing patients receiving baseline S1R agonist SSRIs to those receiving non-S1R agonist SSRIs, no significant decrease in RR was observed. Our findings suggest that use of SSRIs, initiated prior to diagnosis of COVID-19, may be effective in reducing the risk of long-COVID compared with controls. We did not, however, find conclusive evidence to attribute this effect to SSRI-related S1R-agonism, as a comparison of recipients of S1R agonist SSRIs with patients receiving SSRIs that bind either extremely poorly to the S1R (paroxetine) or exert antagonist activity at the S1R (sertraline), did not reveal a significant difference. We speculate that part of the reason why a statistically significant difference was observed for both S1R agonist SSRIs and non-S1R agonist SSRIs versus controls may reside in protective effects unrelated to S1R agonism. We did not see a higher risk of PASC among the group of patients receiving either paroxetine or sertraline even though sertraline is an S1R antagonist. We hypothesize that this is related to the fact that typical free circulating concentrations of sertraline relative to its dissociation constant are low (see Table S1 in the Supplementary Appendix).

As discussed in detail in the introduction, the pathogenesis of PASC has not yet been firmly established, but immune dysregulation with an elevation in proinflammatory cytokine levels may play a role in the condition. We speculate that SSRIs may attenuate such an aberrant response through immunomodulation, as described extensively in the introduction. In addition, SSRIs exert antiplatelet activity, which might, hypothetically, provide benefit in PASC, as microclots may contribute to the pathology of PASC.^16^

Several antidepressant drugs, such as fluoxetine, have been shown to have antiviral properties. The *in vitro* antiviral effect of fluoxetine has been tentatively ascribed to a reduction in intra-lysosomal ceramide related to inhibition of acid sphingomyelinase, but fluoxetine exhibits only low micromolar antiviral potency^31^ raising doubt that it would show evidence of in vivo antiviral efficacy at clinically achieved concentrations. In a prospective randomized double-blind controlled study of COVID-19 outpatients, fluvoxamine exerted no antiviral effect.^40^

Retrospective studies evaluating the potential clinical benefit of SSRIs in hospitalized patients with acute COVID-19 have yielded conflicting findings, ranging from protective effects, such as decreased risk of intubation or death, to absence of any discernible benefit.^45–47^ Similarly, several,^39,40^ though not all^41,42^ prospective, randomized, controlled investigations into the potential therapeutic effects of fluvoxamine in outpatients with acute COVID-19, have suggested evidence of benefit. To our knowledge, this is the first study to suggest a diminished risk of the development of Long COVID in patients receiving SSRIs at baseline. Although an immunological basis for this observation has been postulated, we acknowledge the possibility that the findings in the current study may reflect non-immunological (e.g., antiplatelet or centrally acting) effects of SSRIs. Future prospective studies of SSRIs in patients with PASC or at high risk for PASC that also include immunologic biomarkers seem warranted.

This study has several limitations. First, as a retrospective study using IPW, residual confounding and confounding by indication are two important concerns. Notably, at baseline, the treated group in the main analysis had a higher percentage of female and white individuals and had more comorbidities than the control group, although these differences were mitigated by IPW. For confounding by indication, we attempted to minimize this through the inclusion of other medications that shared indications with SSRIs. Second, the ICD diagnosis code for PASC (U09.9) is of recent vintage, so it is likely that many subjects in N3C without that code may have had PASC. We restricted our analysis to the time period during which the code has been available but, since the U09.9 code was not immediately and widely adopted at all sites, we relied on ML predictions of the PASC phenotype to identify patients more likely to have the condition, including those without a U09.9 code. Using the model’s predictions requires the selection of a probability threshold beyond which patients are labeled with the outcome. We conducted a sensitivity analysis which did establish a degree of robustness to our choice of prediction threshold. Nevertheless, any machine learning model suffers from some degree of bias which may be heterogeneous with respect to data partner, time, and patient records. Additionally, the ML model relies exclusively on structured data, whereas symptoms and signs of PASC may not be fully captured in structured data fields of de-identified EHRs. Although the ML model did not include the medications investigated in this study as predictors, it did include several of the comorbidities we included as covariates (diabetes, chronic kidney disease, congestive heart failure, and chronic lung disease). We do not consider this to be a major concern since (a) IPW balances these characteristics in the exposed and unexposed groups, and (b) all comorbidities of concern are more prevalent in the exposed population (see Table 1).

Another limitation imposed by the ML model is the inability to conduct a sensitivity analysis on patient healthcare interaction. Since N3C contains de-identified EHR records, it is not possible to distinguish between previously healthy patients with a genuine lack of baseline medical history and patients who are new to a healthcare organization for whom prior medical history was not collected. The same applies to interactions with a healthcare organization after the index date: there are many possible reasons why a patient may have no further interactions. Our analyses were therefore limited to patients with some record of interaction with the healthcare organization prior to and after their index diagnosis. Yet since the ML model has a 90 day blackout window around the COVID-19 index date, we could not assess how a lack of baseline medical history and loss to follow-up affected the analysis. We were further unable to study the competing risk of death during the acute phase, defined as within 45 days of the COVID-19 index date. However, patients who died represented a very small fraction of the total population (118/109 183).

Third, data on vaccination, which may affect the risk of PASC, are potentially incompletely captured in the N3C database, though we did mitigate this by selecting sites with vaccination rates matching CDC records. Fourth, our primary analyses did not consider timing, duration, and dosage of the medications at baseline. However, these medications are typically administered chronically and therefore patient exposure will typically be for an extended period of time. We further conducted a sensitivity analysis by restricting treatment to patients who were exposed to the SSRIs within 180 days prior to their COVID-19 index diagnosis date. This analysis is considered to be more rigorous in capturing active SSRI use prior to infection and the resulting estimates remained significant. Fifth, by exclusively relying on de-identified EHR records and selecting for patients with any baseline medical history, the data may not be representative of the general population. Patients with limited access to healthcare or those who seek care at small practices or community hospitals are likely underrepresented. Finally, our results are applicable only to the Delta and Omicron variant given the period of study. Future use of adapted machine learning models to reliably identify additional PASC patients prior to the advent of the U09.9 code may be useful to increase the sizes of the populations in analyses, enabling a finer analysis of the contributions of individual SSRI agents and the influence of specific variants on the risk of PASC.

## Conclusion

In this study, baseline exposure to either S1R agonist or non-S1R agonist SSRIs was associated with a significant reduction in relative risk of long-COVID. Our findings can be used to guide additional research into mechanisms of action of SSRIs on COVID-19. Furthermore, as another contribution to the growing body of evidence on the potential effectiveness of SSRIs as COVID-19 therapeutics, it adds to the need for more definitive double-blind randomized clinical trials.

## Data Availability

The N3C data transfer to NCATS is performed under a Johns Hopkins University Reliance Protocol # IRB00249128 or individual site agreements with NIH. The N3C Data Enclave is managed under the authority of the NIH; information can be found at ncats.nih.gov/n3c/resources. Enclave data is protected, and can be accessed for COVID-related research with an approved (1) IRB protocol and (2) Data Use Request (DUR). Enclave and data access instructions can be found at https://covid.cd2h.org/for-researchers; all code used to produce the analyses in this manuscript is available within the N3C Enclave to users with valid login credentials to support reproducibility.

## Acknowledgements

Authorship was determined using ICMJE recommendations.

We acknowledge Josh Fessel, Leonie Misquitta, and Michael G. Kurilla for valuable discussions and feedback.

We gratefully acknowledge the following core contributors to N3C:

Adam B. Wilcox, Adam M. Lee, Alexis Graves, Alfred (Jerrod) Anzalone, Amin Manna, Amit Saha, Amy Olex, Andrea Zhou, Andrew E. Williams, Andrew Southerland, Anita Walden, Anjali A. Sharathkumar, Benjamin Amor, Benjamin Bates, Brian Hendricks, Brijesh Patel, Caleb Alexander, Carolyn Bramante, Cavin Ward-Caviness, Charisse Madlock-Brown, Christine Suver, Christopher Chute, Christopher Dillon, Chunlei Wu, Clare Schmitt, Cliff Takemoto, Dan Housman, Davera Gabriel, David A. Eichmann, Diego Mazzotti, Don Brown, Eilis Boudreau, Elaine Hill, Elizabeth Zampino, Emily Carlson Marti, Evan French, Farrukh M Koraishy, Federico Mariona, Fred Prior, George Sokos, Greg Martin, Harold Lehmann, Heidi Spratt, Hemalkumar Mehta, Hongfang Liu, J.W. Awori Hayanga, Jami Pincavitch, Jaylyn Clark, Jeremy Richard Harper, Jessica Islam, Jin Ge, Joel Gagnier, Joel H. Saltz, Joel Saltz, Johanna Loomba, John Buse, Jomol Mathew, Joni L. Rutter, Julie A. McMurry, Justin Guinney, Justin Starren, Karen Crowley, Katie Rebecca Bradwell, Kellie M. Walters, Ken Wilkins, Kenrick Dwain Cato, Kimberly Murray, Kristin Kostka, Lavance Northington, Lee Allan Pyles, Leonie Misquitta, Lesley Cottrell, Lili Portilla, Mariam Deacy, Mark M. Bissell, Marshall Clark, Mary Emmett, Mary Morrison Saltz, Matvey B. Palchuk, Melissa A. Haendel, Meredith Adams, Meredith Temple-O’Connor, Michele Morris, Nabeel Qureshi, Nasia Safdar, Nicole Garbarini, Noha Sharafeldin, Ofer Sadan, Patricia A. Francis, Penny Wung Burgoon, Peter Robinson, Philip R.O. Payne, Rafael Fuentes, Randeep Jawa, Rebecca Erwin-Cohen, Rena Patel, Richard A. Moffitt, Richard L. Zhu, Rishi Kamaleswaran, Robert Hurley, Robert T. Miller, Saiju Pyarajan, Samuel Bozzette, Sandeep Mallipattu, Satyanarayana Vedula, Scott Chapman, Shawn T. O’Neil, Soko Setoguchi, Stephanie S. Hong, Steve Johnson, Tellen D. Bennett, Tiffany Callahan, Umit Topaloglu, Usman Sheikh, Valery Gordon, Vignesh Subbian, Warren A. Kibbe, Wenndy Hernandez, Will Beasley, Will Cooper, William Hillegass, Xiaohan Tanner Zhang. Details of contributions available at covid.cd2h.org/core-contributors

## Authors’ contributions

Authorship was determined using ICMJE recommendations.

- Conception: HS, DKS, NH, SGM, KG
- Design: HS, DKS
- Analysis: HS, ATG
- Manuscript drafting: HS, DKS
- Critical revision of manuscript: HS, DKS, ATG, NH, SGM, KG

## The N3C Consortium

Emily R Pfaff; Richard A Moffitt; Christopher G Chute; Melissa A Haendel

## Data sharing statement

The analyses described in this publication were conducted with data or tools accessed through the NCATS N3C Data Enclave covid.cd2h.org/enclave and supported by CD2H - The National COVID Cohort Collaborative (N3C) IDeA CTR Collaboration 3U24TR002306-04S2 NCATS U24 TR002306. This research was possible because of the patients whose information is included within the data from participating organizations (covid.cd2h.org/dtas) and the organizations and scientists (covid.cd2h.org/duas) who have contributed to the on-going development of this community resource. Enclave data is protected, and can be accessed for COVID-related research with an approved (1) IRB protocol and (2) Data Use Request (DUR). Enclave and data access instructions can be found at https://covid.cd2h.org/for-researchers; all code used to produce the analyses in this manuscript is available within the N3C Enclave to users with valid login credentials to support reproducibility.

### Data Partners with Released Data

The following institutions whose data is released or pending:

Available: Advocate Health Care Network — UL1TR002389: The Institute for Translational Medicine (ITM) • Boston University Medical Campus — UL1TR001430: Boston University Clinical and Translational Science Institute • Brown University — U54GM115677: Advance Clinical Translational Research (Advance-CTR) • Carilion Clinic — UL1TR003015: iTHRIV Integrated Translational health Research Institute of Virginia • Charleston Area Medical Center — U54GM104942: West Virginia Clinical and Translational Science Institute (WVCTSI) • Children’s Hospital Colorado — UL1TR002535: Colorado Clinical and Translational Sciences Institute • Columbia University Irving Medical Center — UL1TR001873: Irving Institute for Clinical and Translational Research • Duke University — UL1TR002553: Duke Clinical and Translational Science Institute • George Washington Children’s Research Institute — UL1TR001876: Clinical and Translational Science Institute at Children’s National (CTSA-CN) • George Washington University — UL1TR001876: Clinical and Translational Science Institute at Children’s National (CTSA-CN) • Indiana University School of Medicine — UL1TR002529: Indiana Clinical and Translational Science Institute • Johns Hopkins University — UL1TR003098: Johns Hopkins Institute for Clinical and Translational Research • Loyola Medicine — Loyola University Medical Center • Loyola University Medical Center — UL1TR002389: The Institute for Translational Medicine (ITM) • Maine Medical Center — U54GM115516: Northern New England Clinical & Translational Research (NNE-CTR) Network • Massachusetts General Brigham — UL1TR002541: Harvard Catalyst • Mayo Clinic Rochester UL1TR002377: Mayo Clinic Center for Clinical and Translational Science (CCaTS) • Medical University of South Carolina — UL1TR001450: South Carolina Clinical & Translational Research Institute (SCTR) • Montefiore Medical Center — UL1TR002556: Institute for Clinical and Translational Research at Einstein and Montefiore • Nemours — U54GM104941: Delaware CTR ACCEL Program • NorthShore University HealthSystem — UL1TR002389: The Institute for Translational Medicine (ITM) • Northwestern University at Chicago — UL1TR001422: Northwestern University Clinical and Translational Science Institute (NUCATS) • OCHIN — INV-018455: Bill and Melinda Gates Foundation grant to Sage Bionetworks • Oregon Health & Science University — UL1TR002369: Oregon Clinical and Translational Research Institute • Penn State Health Milton S. Hershey Medical Center — UL1TR002014: Penn State Clinical and Translational Science Institute • Rush University Medical Center — UL1TR002389: The Institute for Translational Medicine (ITM) • Rutgers, The State University of New Jersey — UL1TR003017: New Jersey Alliance for Clinical and Translational Science • Stony Brook University — U24TR002306 • The Ohio State University — UL1TR002733: Center for Clinical and Translational Science • The State University of New York at Buffalo — UL1TR001412: Clinical and Translational Science Institute • The University of Chicago — UL1TR002389: The Institute for Translational Medicine (ITM) • The University of Iowa — UL1TR002537: Institute for Clinical and Translational Science • The University of Miami Leonard M. Miller School of Medicine — UL1TR002736: University of Miami Clinical and Translational Science Institute • The University of Michigan at Ann Arbor — UL1TR002240: Michigan Institute for Clinical and Health Research • The University of Texas Health Science Center at Houston — UL1TR003167: Center for Clinical and Translational Sciences (CCTS) • The University of Texas Medical Branch at Galveston — UL1TR001439: The Institute for Translational Sciences • The University of Utah — UL1TR002538: Uhealth Center for Clinical and Translational Science • Tufts Medical Center — UL1TR002544: Tufts Clinical and Translational Science Institute • Tulane University — UL1TR003096: Center for Clinical and Translational Science • University Medical Center New Orleans — U54GM104940: Louisiana Clinical and Translational Science (LA CaTS) Center • University of Alabama at Birmingham — UL1TR003096: Center for Clinical and Translational Science • University of Arkansas for Medical Sciences — UL1TR003107: UAMS Translational Research Institute • University of Cincinnati — UL1TR001425: Center for Clinical and Translational Science and Training • University of Colorado Denver, Anschutz Medical Campus — UL1TR002535: Colorado Clinical and Translational Sciences Institute • University of Illinois at Chicago — UL1TR002003: UIC Center for Clinical and Translational Science • University of Kansas Medical Center — UL1TR002366: Frontiers: University of Kansas Clinical and Translational Science Institute • University of Kentucky — UL1TR001998: UK Center for Clinical and Translational Science • University of Massachusetts Medical School Worcester — UL1TR001453: The UMass Center for Clinical and Translational Science (UMCCTS) • University of Minnesota — UL1TR002494: Clinical and Translational Science Institute • University of Mississippi Medical Center — U54GM115428: Mississippi Center for Clinical and Translational Research (CCTR) • University of Nebraska Medical Center — U54GM115458: Great Plains IDeA-Clinical & Translational Research • University of North Carolina at Chapel Hill — UL1TR002489: North Carolina Translational and Clinical Science Institute • University of Oklahoma Health Sciences Center — U54GM104938: Oklahoma Clinical and Translational Science Institute (OCTSI) • University of Rochester — UL1TR002001: UR Clinical & Translational Science Institute • University of Southern California — UL1TR001855: The Southern California Clinical and Translational Science Institute (SC CTSI) • University of Vermont — U54GM115516: Northern New England Clinical & Translational Research (NNE-CTR) Network • University of Virginia — UL1TR003015: iTHRIV Integrated Translational health Research Institute of Virginia • University of Washington — UL1TR002319: Institute of Translational Health Sciences • University of Wisconsin-Madison — UL1TR002373: UW Institute for Clinical and Translational Research • Vanderbilt University Medical Center — UL1TR002243: Vanderbilt Institute for Clinical and Translational Research • Virginia Commonwealth University — UL1TR002649: C. Kenneth and Dianne Wright Center for Clinical and Translational Research • Wake Forest University Health Sciences — UL1TR001420: Wake Forest Clinical and Translational Science Institute • Washington University in St. Louis — UL1TR002345: Institute of Clinical and Translational Sciences • Weill Medical College of Cornell University — UL1TR002384: Weill Cornell Medicine Clinical and Translational Science Center • West Virginia University — U54GM104942: West Virginia Clinical and Translational Science Institute (WVCTSI)

Submitted: Icahn School of Medicine at Mount Sinai — UL1TR001433: ConduITS Institute for Translational Sciences • The University of Texas Health Science Center at Tyler — UL1TR003167: Center for Clinical and Translational Sciences (CCTS) • University of California, Davis — UL1TR001860: UCDavis Health Clinical and Translational Science Center • University of California, Irvine — UL1TR001414: The UC Irvine Institute for Clinical and Translational Science (ICTS) • University of California, Los Angeles — UL1TR001881: UCLA Clinical Translational Science Institute • University of California, San Diego — UL1TR001442: Altman Clinical and Translational Research Institute • University of California, San Francisco — UL1TR001872: UCSF Clinical and Translational Science Institute

Pending: Arkansas Children’s Hospital — UL1TR003107: UAMS Translational Research Institute • Baylor College of Medicine — None (Voluntary) • Children’s Hospital of Philadelphia — UL1TR001878: Institute for Translational Medicine and Therapeutics • Cincinnati Children’s Hospital Medical Center — UL1TR001425: Center for Clinical and Translational Science and Training • Emory University — UL1TR002378: Georgia Clinical and Translational Science Alliance • HonorHealth — None (Voluntary) • Loyola University Chicago — UL1TR002389: The Institute for Translational Medicine (ITM) • Medical College of Wisconsin — UL1TR001436: Clinical and Translational Science Institute of Southeast Wisconsin • MedStar Health Research Institute — UL1TR001409: The Georgetown-Howard Universities Center for Clinical and Translational Science (GHUCCTS) • MetroHealth — None (Voluntary) • Montana State University — U54GM115371: American Indian/Alaska Native CTR • NYU Langone Medical Center — UL1TR001445: Langone Health’s Clinical and Translational Science Institute • Ochsner Medical Center — U54GM104940: Louisiana Clinical and Translational Science (LA CaTS) Center • Regenstrief Institute — UL1TR002529: Indiana Clinical and Translational Science Institute • Sanford Research — None (Voluntary) • Stanford University — UL1TR003142: Spectrum: The Stanford Center for Clinical and Translational Research and Education • The Rockefeller University — UL1TR001866: Center for Clinical and Translational Science • The Scripps Research Institute — UL1TR002550: Scripps Research Translational Institute • University of Florida — UL1TR001427: UF Clinical and Translational Science Institute • University of New Mexico Health Sciences Center — UL1TR001449: University of New Mexico Clinical and Translational Science Center • University of Texas Health Science Center at San Antonio — UL1TR002645: Institute for Integration of Medicine and Science • Yale New Haven Hospital — UL1TR001863: Yale Center for Clinical Investigation

